# Genotype-Phenotype Correlations Identify Phenotypic Differences in Sarcomere Mutation-Positive Hypertrophic Cardiomyopathy in the NHLBI HCM Registry

**DOI:** 10.64898/2026.07.28.26359171

**Authors:** Jonathan H. Chan, Christopher Grace, Kate Thomson, Dong-Yun Kim, Patrice Desvigne-Nickens, Paul Kolm, John P. DiMarco, Milind Desai, Raymond Y. Kwong, Carolyn Y. Ho, William Weintraub, Stefan Neubauer, Christopher M. Kramer, Hugh Watkins, Anuj Goel HCMR Investigators

## Abstract

**Background:** Previous studies of genotype-phenotype correlations in hypertrophic cardiomyopathy (HCM) have presented inconsistent conclusions, potentially reflecting small studies and non-uniform phenotyping.

**Objectives:** We aimed to identify genotype-phenotype correlations in sarcomeric variant-positive HCM cardiac magnetic resonance (CMR) imaging and adverse outcome data in the National Heart, Lung, and Blood Institute (NHLBI) HCM Registry.

**Methods:** Of 2750 overall patients, 915 sarcomeric variant-positive ones were subgrouped by genotype. CMR measures of left ventricular (LV) hypertrophy, fibrosis and function, and adverse events, were compared. Findings were meta-analyzed with prior studies from systematic review.

**Results:** Patients with pathogenic variants in thick-filament genes had greater hypertrophy than those in thin-filament genes - maximal LV wall thickness (maxLVWT) (21.7 ± 5.0 vs. 20.0 ± 4.4mm, *P*<0.01) and indexed LV mass (81.9 ± 26.0 vs. 71.7 ± 13.3g/m^2^, *P*<0.001). Of the former, *MYBPC3* carriers had greater maxLVWT than *MYH7* carriers (22.2 ± 5.2 vs. 20.9 ± 4.5mm, *P*<0.01) but lower LV ejection fraction (63.1 ± 8.4 vs. 65.3 ± 8.4%, *P*<0.001). Findings remained significant after covariate adjustment and meta-analysis. 23 (∼1%) patients had >1 disease-linked variants with only 3 (∼0.11%) carrying >1 pathogenic variants. *MYBPC3* carriers had lower risk of a multiple event composite than *MYH7* carriers.

**Conclusions:** In the largest CMR-based study to date, we identified significant phenotypic differences that characterize disease-gene subgroups and resolved prior discrepancies through systematic review and meta-analysis. However, carriage of >1 sarcomeric variants did not contribute much to variation in phenotypic severity in the general HCM population due to its rarity.

**Condensed Abstract:** Prior studies of genotype-phenotype correlations in hypertrophic cardiomyopathy have produced inconsistent findings. In the largest study to date with CMR imaging, we identified significant correlations between disease phenotypes and genotype subgroups of sarcomeric variant-positive patients from the NHLBI Hypertrophic Cardiomyopathy Registry. Patients with disease-linked *MYBPC3* variants had greater hypertrophy and lower ejection fraction than those with *MYH7* variants but also less incidence of clinical outcomes. Patients with multiple variants were strikingly rare (∼1% of all patients). Ultimately, we discovered phenotypic differences that characterize rare genotype subgroups and resolved prior discrepancies through systematic review and meta-analysis.

---

Hypertrophic cardiomyopathy (HCM) is one of the most common inherited heart diseases, affecting 1 in 200-500 people worldwide^1,2^. It is often inherited in an autosomal dominant manner due to rare genetic variants in 8 core genes encoding cardiac sarcomeric proteins. These variants are found in 30-60% of patients^2,3^, indicating the contributing role of other risk factors such as age^4^, sex^5^, common genetic variants^6^, obesity^7^, hypertension^7^ and others^8^. Nevertheless, ‘sarcomere-positive’ patients have 2-fold HCM-related mortality rate than those without such variants^9^. While such studies have presented insightful comparisons between sarcomere-positive and sarcomere-negative HCM^9–11^, how disease expression is influenced by different genotype subgroups within the former remains unclear.

Initial family studies suggested quantitative differences between patients carrying variants from different disease-gene groups, such as less marked hypertrophy in patients with variants in thin filament-encoding genes^12,13^. Functional and modelling studies have also revealed mechanistic differences. For example, variants in thick-filament genes have different mechanistic effects than those in thin-filament genes^14,15^. Recent cohort studies have presented mixed conclusions for genotype-phenotype correlations however^16–20^. This inconsistency may be due to their often single-center nature with small sample sizes limiting statistical power to resolve associations^12,13,16,19^. Discrepancies may also reflect non-uniform phenotyping and selection biases such as recruitment for differing clinical presentations across studies. Past studies have also failed to provide systematic comparisons accounting for multiple testing burden or confounding factors such as age and sex^17–21^.

The National Heart, Lung and Blood Institute (NHLBI) HCM Registry (HCMR) is the first large prospective registry of HCM patients (N = 2750) to combine cardiac magnetic resonance (CMR) imaging and genetic testing at baseline, with clinical follow-up^1,11^. CMR is regarded as the gold-standard for phenotyping of cardiac structure and function in HCM^22^, and provides measures of left ventricular (LV) hypertrophy, fibrosis, and function. Such imaging-derived phenotypes reflect HCM pathobiology and serve as measures of disease severity in conjunction with adverse clinical outcome data. We applied *a priori* approaches to subgroup 915 sarcomere-positive patients by their genotype and compared phenotypes across these distinct subgroups. We further meta-analyzed cardiac imaging-derived phenotype associations from HCMR with prior studies identified by systematic review to increase statistical power and resolve existing discrepancies. Thus, we determined genotype-phenotype correlations in sarcomere-positive HCM in the largest CMR-based study to date.

## Methods

### Hypertrophic Cardiomyopathy Registry

The Hypertrophic Cardiomyopathy Registry (HCMR) is a prospective observational study of 2750 HCM patients enrolled across 44 centers in 6 countries across North America and Europe from April 1, 2014 to April 7, 2017^1,11,23^. All patients provided written informed consent and were subject to clinical evaluation including imaging (echocardiography and CMR) and genetic analyses. The study population, inclusion and exclusion criteria have been previously detailed^11^. The National Research Ethics Service approved the HCMR study (14/SC/0190).

CMR imaging was performed via 1.5T or 3.0T systems (General Electric, Philips Medical Systems or Siemens Healthineers) using a standardized protocol (Supplemental Methods 1)^11^. Patients were also sequenced at 36 cardiomyopathy-associated genes, including the core 8 sarcomeric genes causally linked to HCM *(MYBPC3*, *MYH7*, *MYL2*, *MYL3*, *ACTC1*, *TNNT2*, *TNNI3*, *TPM1*)^3^ using Illumina MiSeq. Downstream analysis and clinical pathogenicity classification according to American College of Medical Genetics/Association for Molecular Pathology guidelines^24^ ensued to classify patients with disease-linked rare variants in these core 8 genes as ‘sarcomere-positive’ or ‘sarcomere-negative’ if otherwise (Supplemental Methods 2)^11^.

### Statistical Analysis

Quality control was performed to remove individuals with missing information. Sarcomere-negative individuals were also excluded (Figure 1).

**Figure 1.**
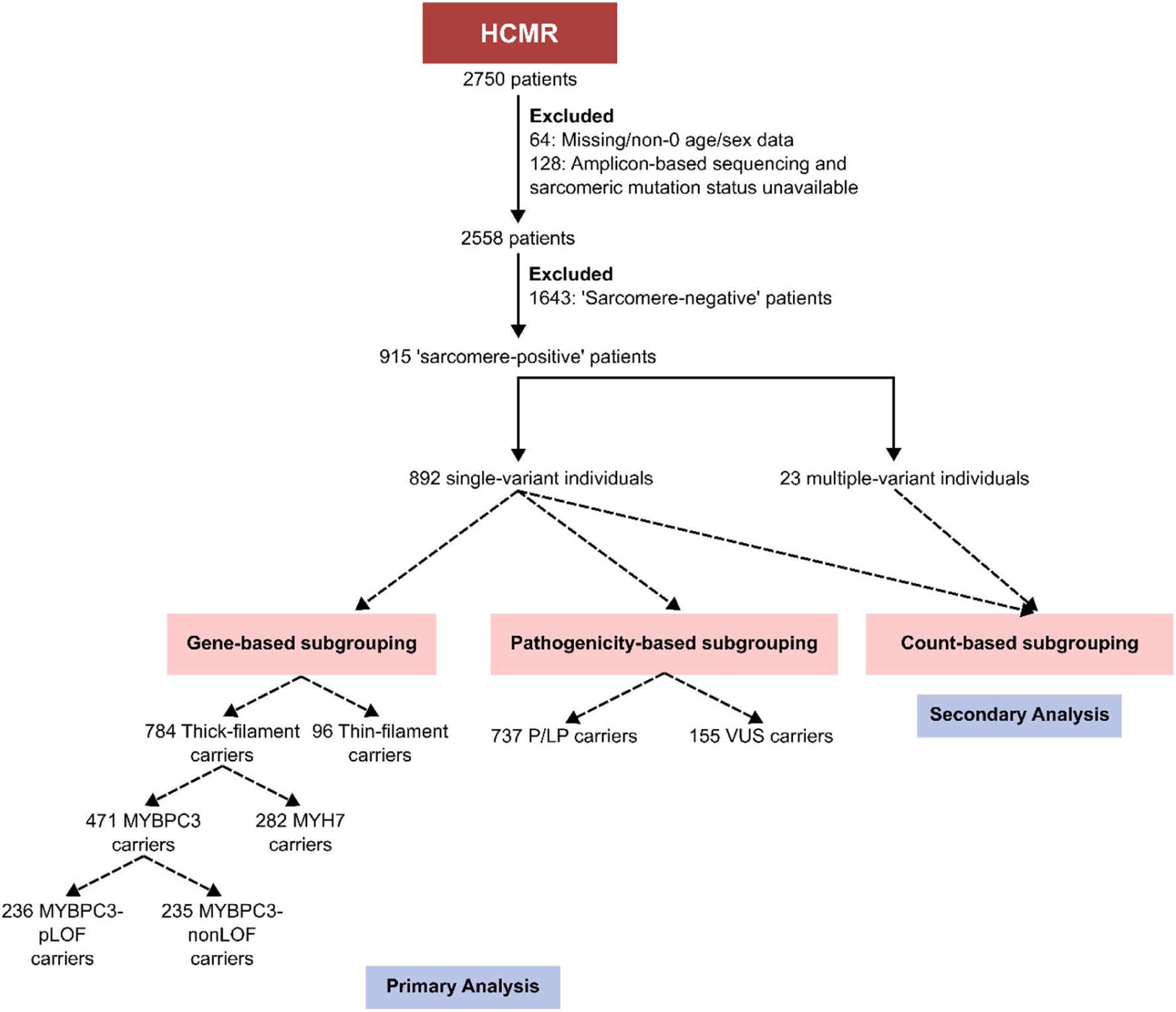
Study flowchart. 2750 patients from the Hypertrophic Cardiomyopathy Registry (HCMR) were filtered for those with rare genetic variants in the core 8 sarcomeric genes (*MYBPC3*, *MYH7*, *MYL2*, *MYL3*, *TNNT2*, *TNNI3*, *TPM1*, *ACTC1*). Individuals with disease-linked variants as per ACMG/AMP^24^ guidelines were labelled ‘sarcomere-positive’^11^. These patients were dichotomised into single-variant individuals or multiple-variant individuals. In primary analysis, the former set were analysed via gene-based and pathogenicity-based subgrouping approaches. In secondary analysis, the multiple-variant patients were compared to the single-variant patients. Solid and dashed arrows indicate filtering and subgrouping of individuals respectively. CMR = cardiac magnetic resonance; P/LP = pathogenic/likely pathogenic; VUS = variant of uncertain significance.

#### Patient Subgrouping

Subgrouping of patients by genotype was based on *a priori* hypotheses involving the gene carrying the rare variant (gene-based subgrouping), clinical pathogenicity of rare variant (pathogenicity-based subgrouping), and number of disease-linked variants carried (count-based subgrouping). For each approach, per-subgroup counts are detailed in Supplemental Tables 1, 2 and 3.

For primary analysis (gene-based and pathogenicity-based approaches), only individuals with a single disease-linked variant were included (Figure 1). Gene-based subgrouping involved grouping single-variant patients by the gene of their disease-linked variant (Figure 1). The following comparisons were made: 1) thick-filament encoding genes (*MYBPC3*, *MYH7*, *MYL2*, *MYL3*) vs. thin-filament encoding genes *(TNNT2*, *TNNI3*, *TPM1*); 2) *MYBPC3* vs. *MYH7*; 3) *MYBPC3*-pLoF (predicted-loss-of-function) vs. *MYBPC3*-nonLoF (non-loss-of-function). *ACTC1* was excluded from the thin-filament subgrouping in comparison 1) as the functional and biochemical properties of *ACTC1* HCM variants differ from those of variants in the thin-filament genes encoding the Ca^2+^-sensitive regulatory troponin complex, with such *ACTC1* variants instead directly impacting actomyosin interactions^25,26^. For comparison 3), the dichotomization of *MYBPC3* variants used VEP (v110)-annotated consequence with disease-linked variants grouped into pLoF if annotated as a stop-gained, frameshift, splice donor or acceptor variant, and predicted as a high-confidence LoF variant by LOFTEE^27^. The NMD plugin was also used to exclude stop-gained variants predicted to escape nonsense-mediated decay (NMD). Such NMD-escaping variants, LOFTEE-failing variants, and other disease-linked *MYBPC3* variants (e.g. missense variants) were grouped into *MYBPC3*-nonLoF. Pathogenicity-based subgrouping involved subgrouping patients by the classified pathogenicity of their rare variant (pathogenic or likely pathogenic (P/LP) or variant of uncertain significance (VUS)).

Secondary analysis applied count-based subgrouping to all sarcomere-positive individuals to compare those with a single disease-linked variant against those with >1 variant (multiple-variant) whether as homozygotes or as compound or double heterozygotes (Figure 1).

#### Phenotype Selection

Of the many phenotypes measured in HCMR^11^, a select few were chosen to minimize multiple testing burden. These included maximal LV wall thickness (maxLVWT) and indexed LV mass (LVMi) reflecting hypertrophy; LGE and ECVF measuring replacement and interstitial fibrosis respectively; LV ejection fraction (LVEF) and median global transmural longitudinal, circumferential, and radial LV strains reflecting function. Sensitivity analyses were also conducted with echocardiography-based measures for maxLVWT and LVEF.

#### Adverse Clinical Outcome Composites

To increase statistical power for time-to-event analyses, clinical events were combined into predefined composites^28^. A primary composite endpoint was defined as first occurrence of cardiovascular death, ventricular tachyarrhythmia (resuscitated cardiac arrest; sustained ventricular tachycardia and appropriate ICD discharge), heart transplantation or LVAD placement. A secondary composite endpoint was defined as first occurrence of primary composite events, as well as non-cardiac death, hospitalization for heart failure (HF), atrial fibrillation (AF) requiring therapy and non-fatal stroke. Sensitivity analyses were conducted using event-focused composites of HF (HF-mediated death, heart transplantation or LVAD placement and hospitalization for HF), AF, and ventricular arrythmias (VA) (arrhythmic cardiac death or ventricular tachyarrhythmia).

#### Statistical Methods

Summary statistics for continuous variables were expressed as mean ± standard deviation (if normally distributed) or median (1^st^ quartile – 3^rd^ quartile) (if non-normally distributed). Pairwise comparisons between subgroups were made via multivariable linear regression to adjust for age, sex, body mass index, MRI field strength, and for non-indexed, non-percentage phenotypes such as maxLVWT, body surface area. Skewed phenotypes were log-transformed for regression analysis. Regression effect size was reported as beta estimate ± standard error (for text) but to enable comparison among the different phenotypes (with varying magnitudes), regression analyses were repeated after standardisation of the phenotype (for plotting). Association with incidence of clinical events was performed via Cox regression, using age as the time scale (to reduce potential confounding^29^) with individuals left-truncated at age of recruitment (delayed-entry to mitigate immortal time bias^9^) and right-censored at the date of study discontinuation, the last follow-up date, or the date of death (if the cause was unknown). Covariates included sex and body mass index. Proportional hazards assumption was evaluated via scaled Schoenfeld residuals plot and significance test, with introduction of a time-varying covariate effect if the assumption was invalid for any predictor. Multiple testing correction for all analyses was applied via Benjamini-Hochberg procedure to control Type I error rate at 5%. Statistical analysis was performed at core laboratory (University of Oxford, Oxford, United Kingdom) via R (v4.3.3).

#### Systematic Review and Meta-Analyses

Systematic review was conducted using PRISMA (Preferred Reporting Items for Systematic Reviews and Meta-Analysis) criteria (Supplemental Figure 1)^30^. The objective was to evaluate cardiac imaging-derived phenotypes across the genotype subgroups of HCM patients with disease-linked rare variants in the core 8 sarcomeric genes. A systematic search of PubMed, Scopus and Web of Science was performed to identify relevant studies published from 2012 (release of population whole-exome/genome sequencing databases as 1000 Genomes Project and Exome Sequencing Project which enabled variant reclassification) to February 23, 2026. Further details of search, study inclusion and exclusion criteria are described in Supplemental Methods 3. Quality assessment of studies was conducted using the National Institutes of Health quality assessment tool for cross-sectional, observational cohort studies. Studies were excluded from meta-analysis if ‘Poor’ quality rating. Meta-analysis was performed using fixed-effects inverse-variance weighted method with mean-difference effect size metric in R v4.3.3 with *meta* R package v7.0 at core laboratory (University of Oxford, Oxford, United Kingdom). Further details are described in Supplemental Methods 4.

## Results

### Baseline Characteristics

Of the 2750 patients enrolled who had not withdrawn consent, 915 were classified as sarcomere-positive, as described in Methods and previously published^11^. Briefly, these were carriers of P/LP and certain VUSs in the core 8 sarcomeric genes. Of the 915 sarcomere-positive subjects, 892 had 1 disease-linked rare variant (Figure 1). Their baseline demographic and clinical information are outlined in Table 1. CMR-derived phenotypes reflecting hypertrophy (maxLVWT and LVMi), fibrosis (LGE and ECVF) and function (LVEF and LV strains) were compared across subgroups with summary statistics detailed in Table 2.

**Table 1.**
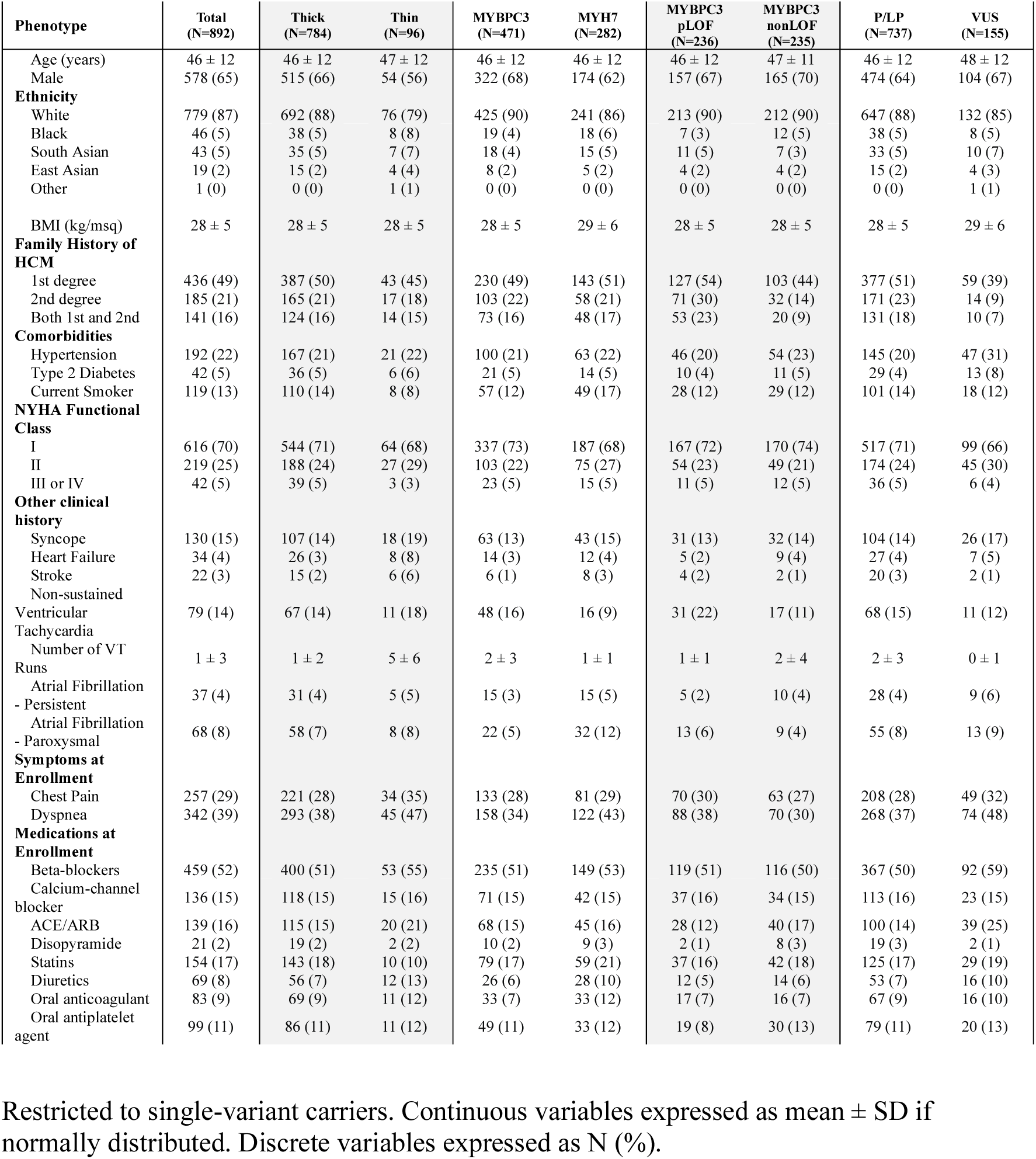
^1^Baseline characteristics of sarcomeric rare variant carriers in Hypertrophic Cardiomyopathy Registry. ACE/ARB = angiotensin-converting enzyme inhibitors/ angiotensin receptor blockers; BMI = body mass index; HCM = hypertrophic cardiomyopathy; nonLOF = non-loss-of-function; NYHA = New York Heart Association; pLOF = predicted loss-of-function; P/LP = pathogenic/likely pathogenic; VT = ventricular tachycardia; VUS = variant of uncertain significance.

**Table 2.** ^2^Cardiac magnetic resonance imaging-derived phenotypes of sarcomeric rare variant carriers in Hypertrophic Cardiomyopathy Registry. LGE = late gadolinium enhancement; LV = left ventricle; nonLOF = non-loss-of-function; pLOF = predicted loss-of-function; P/LP = pathogenic/likely pathogenic; VUS = variant of uncertain significance.

| Phenotype | Total<br>(N=892) | Thick<br>(N=784) | Thin<br>(N=96) | p-value | MYBPC3<br>(N=471) | MYH7<br>(N=282) | p-value | MYBPC3<br>pLOF<br>(N=236) | MYBPC3<br>nonLOF<br>(N=235) | p-value | P/LP<br>(N=737) | VUS<br>(N=155) | p-value |
| --- | --- | --- | --- | --- | --- | --- | --- | --- | --- | --- | --- | --- | --- |
| **Hypertrophy** |  |  |  |  |  |  |  |  |  |  |  |  |  |
| Maximal LV Wall Thickness (mm) | 21.5 ± 5 | 21.7 ± 5 | 20 ± 4.4 | **0.0029** | 22.2 ± 5.2 | 20.9 ± 4.5 | **0.0054** | 22.5 ± 5.5 | 21.9 ± 4.9 | 0.38 | 21.6 ± 5 | 21.3 ± 5.2 | 0.32 |
| LV Mass Index (g/m<sup>2</sup>) | 80.8 ± 25.1 | 81.9 ± 26 | 71.7 ± 13.3 | **0.0009** | 82.7 ± 26.2 | 80.4 ± 25.4 | 0.28 | 84.6 ± 29 | 80.7 ± 22.9 | 0.39 | 80.7 ± 25.7 | 82.3 ± 22.9 | 0.19 |
| **Fibrosis** |  |  |  |  |  |  |  |  |  |  |  |  |  |
| LGE (6SD) (% of LV Mass) | 0.9 (0-3.0) | 0.94 (0-3.1) | 0.85 (0-2.7) | 0.42 | 1.1 (0-4.0) | 0.6 (0-2.1) | **0.003** | 1.07 (0-3.5) | 1.28 (0.1-4.1) | 0.32 | 1.0 (0.01-3.2) | 0.4 (0-2.2) | **0.0017** |
| LV Extracellular Volume Fraction (%) | 31.4 ± 5.1 | 31.3 ± 5.1 | 32.2 ± 4.7 | 0.075 | 31.3 ± 5.2 | 31.3 ± 5.1 | 0.65 | 31.3 ± 4.8 | 31.3 ± 5.6 | 0.71 | 31.5 ± 5.1 | 30.6 ± 5 | **0.037** |
| **Function** |  |  |  |  |  |  |  |  |  |  |  |  |  |
| LV Ejection Fraction (%) | 64 ± 8.4 | 64 ± 8.4 | 64.5 ± 8.5 | 0.54 | 63.1 ± 8.4 | 65.3 ± 8.4 | **0.001** | 63.7 ± 8.6 | 62.5 ± 8.2 | 0.085 | 63.9 ± 8.5 | 64.4 ± 8.3 | 0.56 |
| - Global Longitudinal LV Strain (Transmural) (%) | 11.1 ± 2.5 | 11.1 ± 2.5 | 11.2 ± 2.5 | 0.65 | 10.9 ± 2.5 | 11.3 ± 2.6 | 0.057 | 11.1 ± 2.6 | 10.8 ± 2.4 | 0.45 | 11.1 ± 2.6 | 11 ± 2.4 | 0.74 |
| - Global Circumferential LV Strain (Transmural) (%) | 11.8 ± 2.4 | 11.8 ± 2.5 | 12.2 ± 2.2 | 0.12 | 11.6 ± 2.4 | 12.1 ± 2.4 | **0.014** | 11.6 ± 2.3 | 11.5 ± 2.5 | 0.78 | 11.8 ± 2.5 | 11.7 ± 2.5 | 0.76 |
| Global Radial LV Strain (Transmural) (%) | 33.4 ± 16.6 | 33.1 ± 16.5 | 35.8 ± 16.7 | 0.16 | 32.3 ± 15.7 | 34.2 ± 17.4 | 0.27 | 32.6 ± 15.7 | 32 ± 15.8 | 0.6 | 33.2 ± 16.6 | 33.6 ± 16.4 | 0.76 |

Restricted to single-variant carriers. Continuous variables expressed as mean ± SD if normally distributed or median (1<sup>st</sup>- 3<sup>rd</sup> quartiles) if non-normally distributed. Discrete variables expressed as N (%). P-values derived via 2-sided Wilcoxon Rank Sum test prior to multiple-testing correction and **bold** if nominally significant.

### Gene-based Subgrouping

#### Thick-filament vs. Thin-filament

Prior family and cohort studies indicated phenotypic differences between patients carrying disease-linked rare variants in thick-filament genes, and those carrying variants in thin-filament genes^12,13,17^. In HCMR, the former had significantly greater maxLVWT (21.7 ± 5.0 vs. 20.0 ± 4.4mm, *P*<0.01) and LVMi (81.9 ± 26.0 vs. 71.7 ± 13.3g/m^2^, *P*<0.001) than the latter (Table 2). Both remained significant after covariate adjustment, with the thick-filament subgroup having a 1.5 ± 0.5mm greater maxLVWT and 8.5 ± 2.6g/m^2^ greater LVMi on average compared to the thin-filament one (Figure 2, Supplemental Table 4). Sensitivity analysis using echocardiography-derived maxLVWT further supported this finding (Supplemental Figure 2).

**Figure 2.**
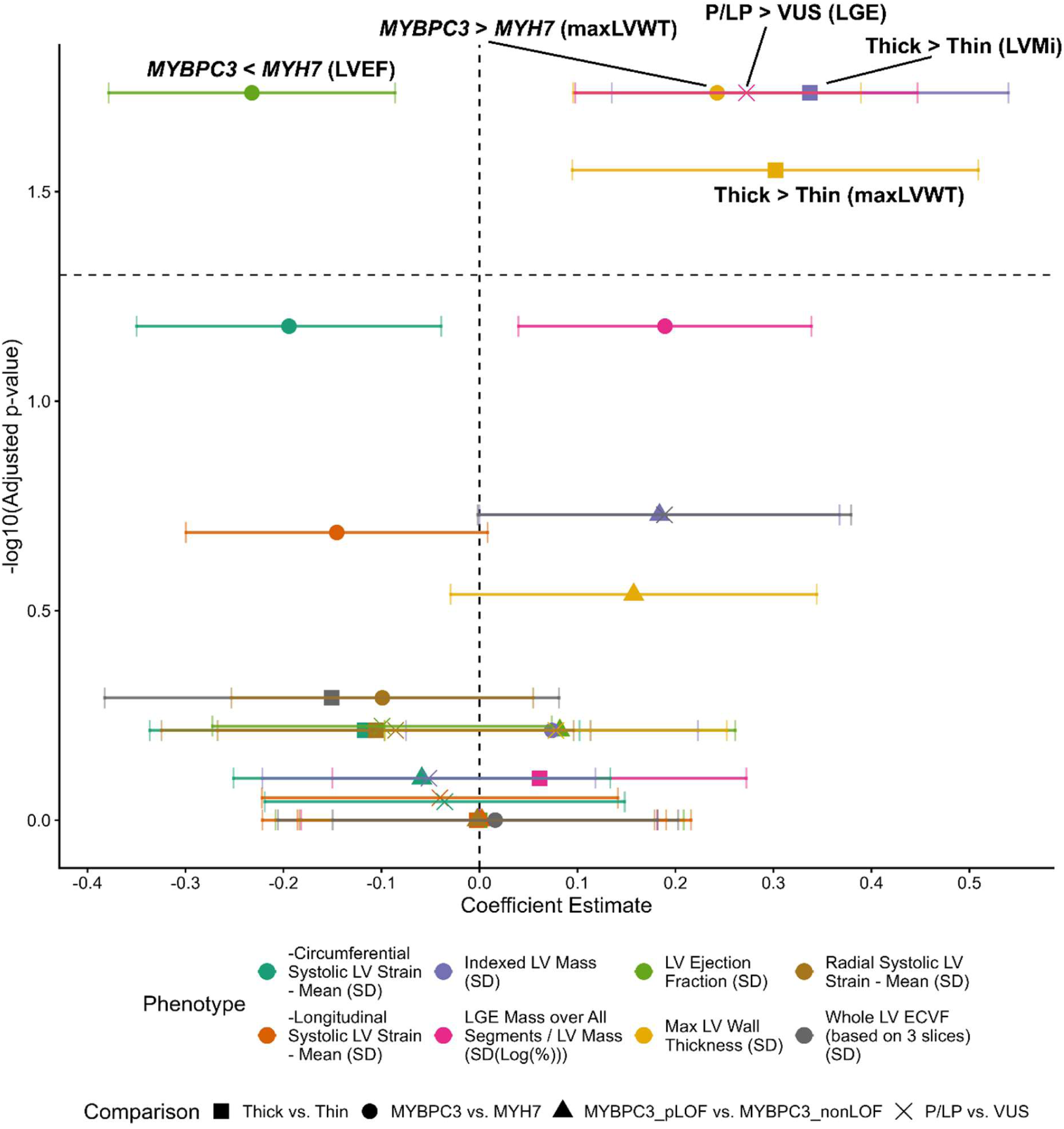
Summary of CMR phenotype comparisons across subgroups via multivariable linear regression. Comparisons were adjusted for covariates of age, sex, body mass index, MRI field strength (and body surface area for maximal LV wall thickness (maxLVWT). Phenotypes were normalised to standard deviation units via scaling to enable comparison across varied scales of magnitude. Statistical test of coefficient inequality from 0 was conducted via t-test with adjustment by Benjamini-Hochberg procedure. Dashed horizontal line indicates 5% adjusted p-value threshold. Significant genotype-phenotype correlations are labelled. Error bars indicate 95% confidence interval. Coefficient estimate reported for first group in each comparison relative to the other and detailed in Supplemental Table 4. ECVF = extracellular volume fraction; LGE = late gadolinium enhancement; LV = left ventricular; nonLOF = non-loss-of-function; pLOF = predicted loss-of-function; P/LP = pathogenic/likely pathogenic; VUS = variant of uncertain significance.

#### *MYBPC3* vs. *MYH7*

Rare genetic variants in *MYBPC3* and *MYH7* constitute the majority of HCM-linked variants in the core 8 sarcomeric genes^2^. Despite both genes encoding thick-filament components, prior studies indicated significant differences including lower contractility^19^ in patients with disease-linked *MYBPC3* variants relative to *MYH7*. Contrastingly, other studies found none^16,18^. In HCMR, *MYBPC3*-positive patients had significantly greater maxLVWT (22.2 ± 5.2 vs. 20.9 ± 4.5mm, *P*<0.01), lower LVEF (63.1 ± 8.4 vs. 65.3 ± 8.4%, *P*<0.001) and lower circumferential strain (11.6 ± 2.4 vs. 12.1 ± 2.4%, *P*<0.05), as well as greater LGE (1.1 (0.0-4.0) vs. 0.6 (0-2.1)%, *P*<0.001) than *MYH7*-positive ones (Table 2). After covariate adjustment, only the maxLVWT and LVEF associations remained significant. Specifically, *MYBPC3*-positive patients had a 1.2 ± 0.4mm greater maxLVWT and a 2.0 ± 0.6% lower LVEF than *MYH7*-positive patients on average (Figure 2, Supplemental Table 4). Sensitivity analyses using echocardiography-derived metrics likewise supported greater hypertrophy and lower LVEF in *MYBPC3*-positive relative to *MYH7*-positive patients (Supplemental Figure 2).

#### *MYBPC3* predicted-LoF vs. *MYBPC3* non-LoF

Within *MYBPC3*-positive patients, both predicted LoF variants and non-LoF variants can cause disease. Mechanistically, they behave differently with the former causing haploinsufficiency^31,32^ and the latter having some which do so and others with dominant-negative effects^32^. This might underlie phenotypic differences but in HCMR, no significant differences were observed (Table 2), including after covariate adjustment (Figure 2).

### Pathogenicity-based Subgrouping

The clinical pathogenicity of disease-linked variants relies on human genetic, experimental, and computational evidence to reflect variants’ penetrance and expressivity^24^. As such, different degrees of pathogenicity affect disease manifestation^10^ and to investigate this, single-variant patients were subgrouped based on their variant’s clinical pathogenicity. Those carrying P/LP variants had significantly greater LGE (1.0 (0.0-3.2) vs. 0.4 (0-2.2) %, *P*<0.01) and ECVF (31.5 ± 5.1 vs. 30.6 ± 5.0%, *P*<0.05) than those carrying VUSs (Table 2). The former signal remained significant after covariate adjustment, with a difference of 0.9 ± 0.5% (Figure 2, Supplemental Table 4). This indicates greater replacement fibrosis associated with P/LP variants than VUSs.

### Meta-analyses

Systematic search identified 70 full texts of which 20 passed inclusion criteria (Supplemental Figure 1 & Supplemental Table 5). Only 10 of these used CMR to compare patient subgroups and most were relatively underpowered, single-center studies^16,19^. Nevertheless, meta-analyses summarized prior findings to reduce uncertainty and deflate potentially inflated effect estimates from HCMR analyses.

For meta-analysis of thick-filament variant-positive patients vs. thin-filament ones, no equivalent prior comparisons of CMR-derived maxLVWT were identified, but meta-analysis of CMR-derived LVMi^17^ (Figure 3a) supported greater hypertrophy in the former. HCMR analyses also showed that *MYBPC3*-positive patients had greater maxLVWT and lower LVEF than *MYH7*-positive patients. Meta-analysis showed that these findings remained significant (Figure 3b)^16,18,19^. Corresponding meta-analyses with prior echocardiography-based studies were also supportive (Supplemental Figure 3b). Meta-analyses with prior studies comparing P/LP-positive patients with VUS-positive patients^21,33^ supported the HCMR finding of greater LGE in the former (Supplemental Figure 5) as well as the lack of other significant phenotypic differences (Supplemental Figure 6c).

**Figure 3.**
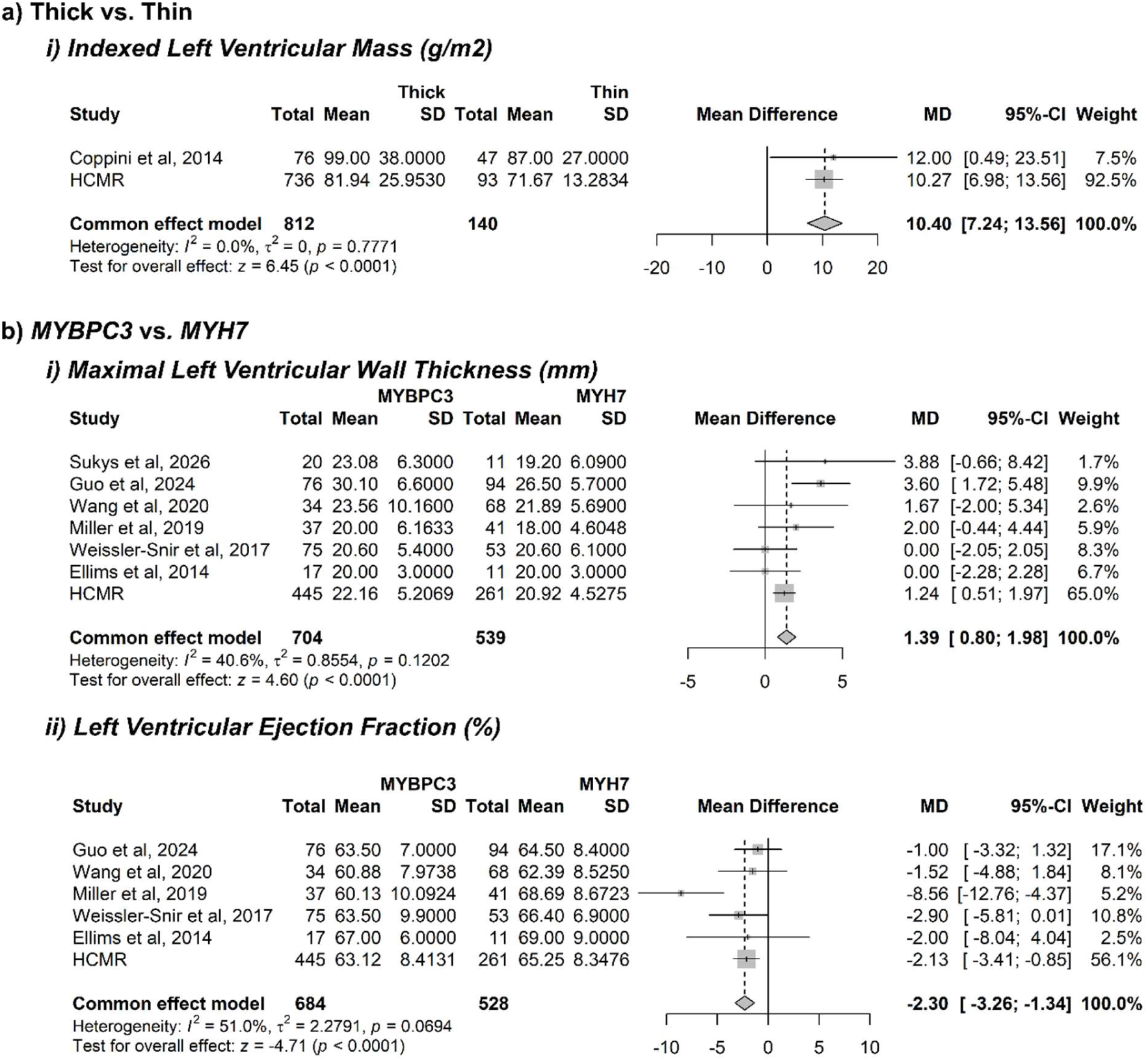
Meta-analyses of significant conclusions from primary analyses of Thick vs. Thin & *MYBPC3* vs. *MYH7* rare variant carriers in HCMR, with prior literature. *ai)* Indexed left ventricular (LV) mass index primary analyses were meta-analysed with prior studies comparing patients with disease-linked rare variants in thick filament-encoding genes vs. thin-filament encoding genes. *bi)* Maximal LV wall thickness and *bii)* LV ejection fraction primary analyses were meta-analysed with prior studies comparing patients with disease-linked rare variants in *MYBPC3* and *MYH7*^16,18,19,46^. Primary analysis conclusions remained significant for, defining these genotype-phenotype correlations of greater maxLVWT and less LVEF in *MYBPC3* carriers relative to *MYH7* carriers. Heterogeneity in *bii)* was addressed via random-effects and leave-one-out analyses (Supplemental Figure 4) suggesting that one study^19^ behaved as an outlier. Meta-analyses were conducted via fixed-effects inverse-variance weighted method on mean-difference effect size metric. CI = confidence interval; HCMR = Hypertrophic Cardiomyopathy Registry; MD = mean-difference; SD = standard deviation.

While some previous studies presented results contrary to HCMR such as thick-filament-positive patients having greater LVEF than thin-filament-positive ones^17^, meta-analysis with HCMR findings did not change the significance of any primary analyses, except for supporting greater LVMi in *MYBPC3*-positive vs. *MYH7*-positive patients (Supplemental Figure 6). This aligns with maxLVWT findings and the consensus of elevated hypertrophy in the former subgroup.

### Count-based Subgrouping

Prior studies have indicated a dosage-dependent effect of disease-linked rare variants to HCM severity^10,34,35^ but this was contested by a more recent systematic review^36^. We investigated this via secondary analysis in HCMR by comparing patients carrying a single disease-linked rare variant with those carrying multiple. Their baseline characteristics are detailed in Supplemental Table 6. First and foremost, such multiple variant carriers were rare (N=23) corresponding to ∼1% of all patients, mostly carried a single P/LP and a VUS variant (N=16/23). 3 patients were compound or double heterozygotes for P/LP variants (Supplemental Table 3), corresponding to ∼0.11% of all patients. Phenotypically, comparing multiple variant carriers to single variant ones revealed no significant phenotypic differences except for greater LGE in the former (3.6 (0.7-7.2) vs. 0.9 (0-3.0) %, *P*<0.001) (Table 4). This remained significant after covariate adjustment and multiple testing correction with 2.7 ± 1.0% greater LGE on average (Figure 4, Supplemental Table 7). The 3 compound/double heterozygotes for P/LP variants also did not appear substantially different in phenotype (Supplemental Table 8). However, such analyses were limited in power by the inherent rarity of multiple variant carriers. Ultimately, this work highlights how carriage of multiple disease-linked variants is rare in HCM and hence not a major contributor to phenotypic heterogeneity.

**Figure 4.**
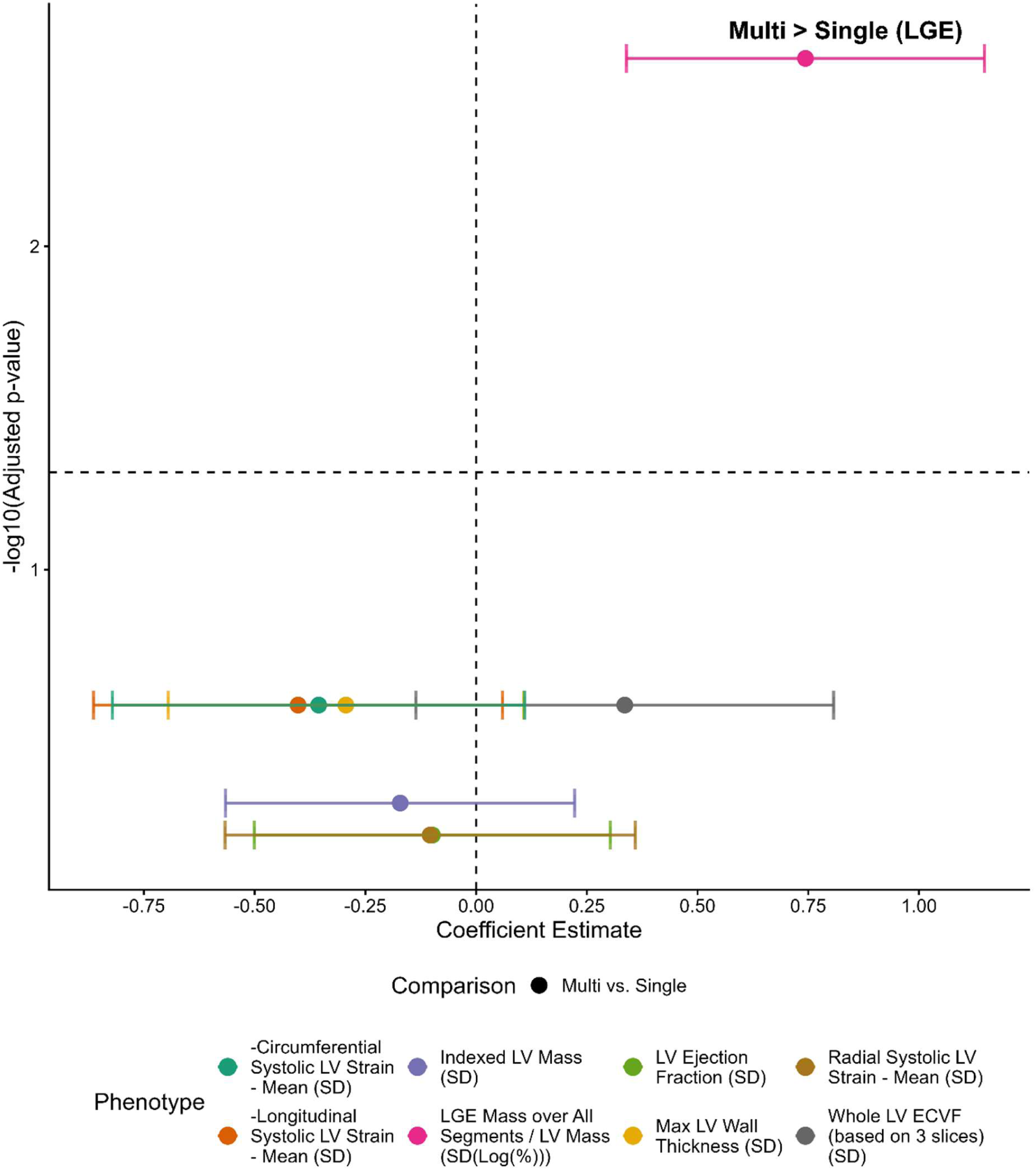
Summary of CMR phenotype comparisons across single vs. multiple-variant sarcomere-positive HCM patients via multivariable linear regression. Comparisons were adjusted for covariates of age, sex, body mass index, MRI field strength, (and body surface area for maximal LV wall thickness (maxLVWT). Phenotypes were standardised to standard deviation units to enable comparison across varied scales of magnitude. Statistical test of coefficient inequality from 0 was conducted via t-test with adjustment by Benjamini-Hochberg procedure. Only significant difference was greater LGE in multiple-variant relative to single-variant patients. Dashed horizontal line indicates 5% adjusted p-value threshold. Significant genotype-phenotype correlations are labelled. Error bars indicate 95% confidence interval. Coefficient estimate reported for first group in each comparison relative to the other and detailed in Supplemental Table 7. ECVF = extracellular volume fraction; LGE = late gadolinium enhancement; LV = left ventricular; nonLOF = non-loss-of-function; pLOF = predicted loss-of-function; P/LP = pathogenic/likely pathogenic; VUS = variant of uncertain significance.

### Adverse Clinical Outcomes

Sarcomere-positive patients were followed-up for a median of 7.6 years [6.2 – 8.1 IQR] with 38 primary composite events and with 149 secondary composite events (Supplemental Table 9). As with imaging-derived phenotypes, incidence of clinical composite outcomes was compared between genetic subgroups. From primary analysis within single-variant individuals, the only significant difference was a substantially lower incidence of the secondary composite endpoint in *MYBPC3*-positive patients relative to *MYH7*-positive ones (Figure 5) with hazard ratio of 0.6 ([0.42 – 0.87] 95% CI) (Table 3). This is illustrated by Kaplan-Meier survival curves (Supplemental Figure 7, Supplemental Figure 8). Sensitivity analysis with event-focused composites showed that this was largely driven by lower incidence of AF but also suggested a trend towards lower HF incidence (Supplemental Figure 9). Of the secondary analysis comparison of single vs. multiple-variant individuals, there were no significant differences in incidence of either primary or secondary composite (Supplemental Table 10).

**Figure 5.**
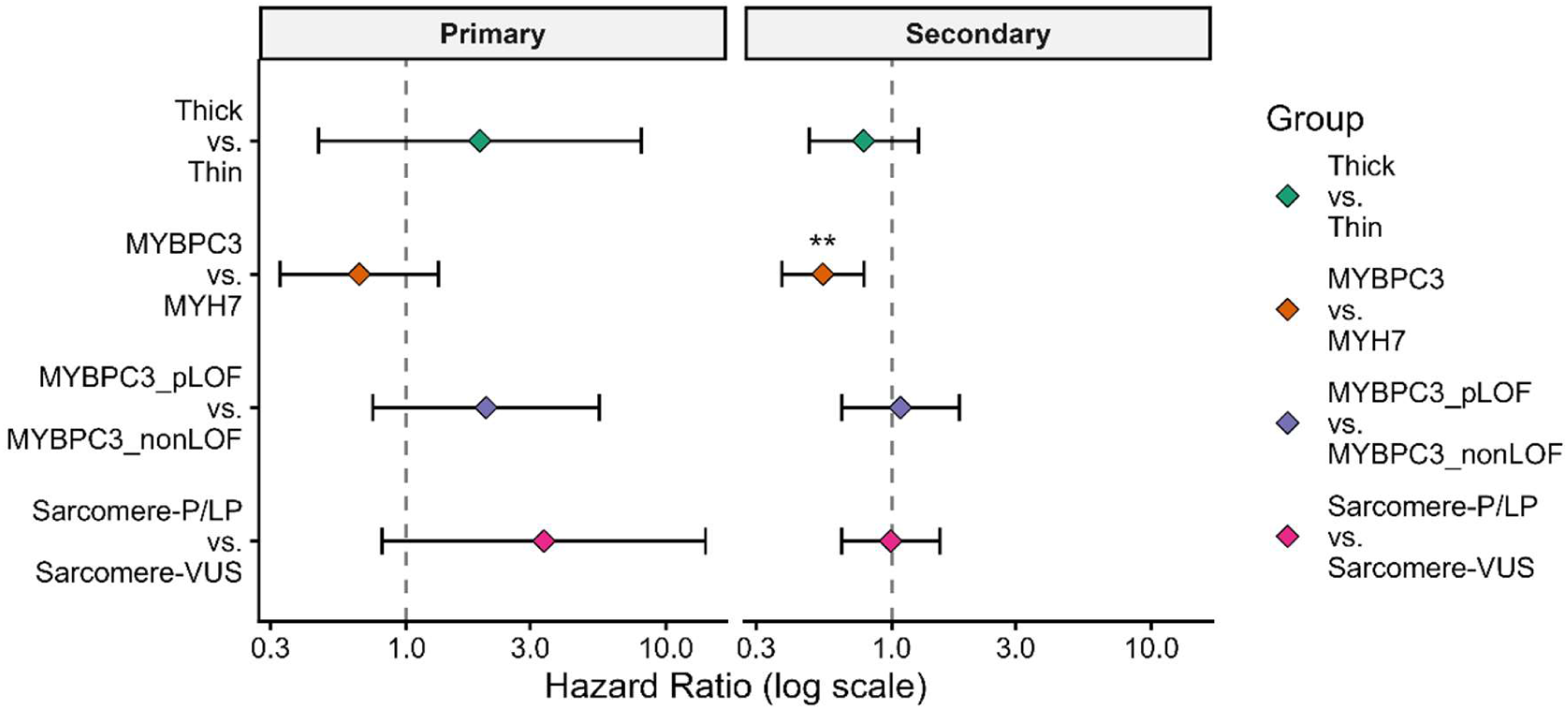
Summary of adverse clinical outcome associations across subgroups via multivariable Cox regression. Patients with disease-linked *MYBPC3* variants had less incidence of the secondary composite outcome relative to those with disease-linked *MYH7* variants. Primary composite included HCM-related cardiac death, ventricular tachyarrhythmia, heart transplantation and left ventricular assist device placement. Secondary composite included the primary events as well as non-cardiac death, hospitalisation for heart failure, atrial fibrillation requiring therapy and non-fatal stroke. Events left-truncated at recruitment date and right-censored at date of study discontinuation, dataset version date or death date (if unknown cause). Associations were adjusted for covariates of sex and body mass index. Statistical test of coefficient inequality from 0 conducted via Wald test with adjustment by Benjamini-Hochberg procedure. Statistical significance indicated by * (adjusted p-value < 0.05). Error bars indicate 95% confidence interval. nonLOF = non-loss-of-function; pLOF = predicted loss-of-function; P/LP = pathogenic/likely pathogenic; VUS = variant of uncertain significance.

**Table 3.**
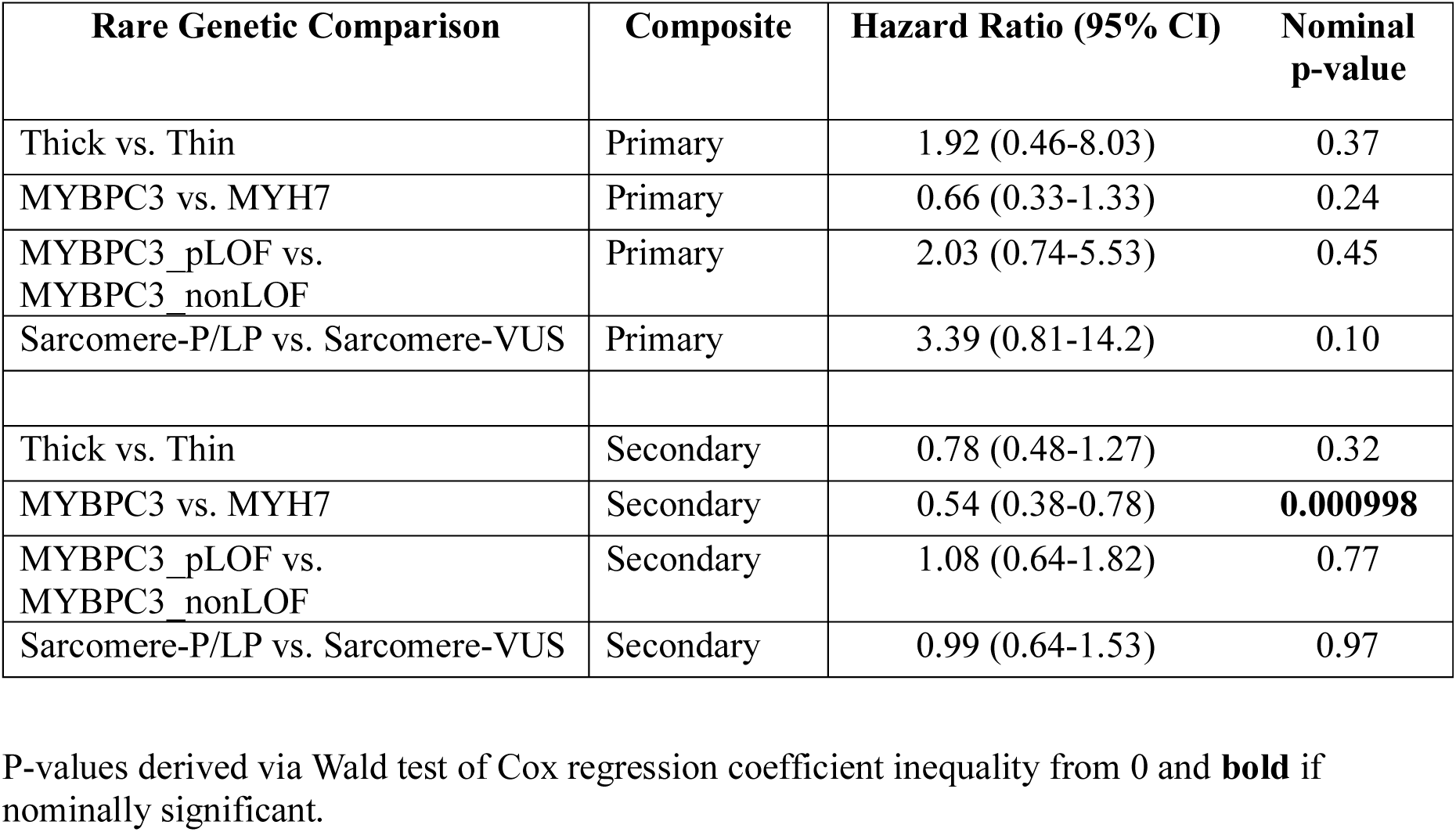
^3^Hazard Ratios from Multivariable Cox Regression of Composite Outcomes in Single-Variant Sarcomere Positive HCMR Patients Across Rare Genetic Subgroups. nonLOF = non-loss-of-function; pLOF = predicted loss-of-function; P/LP = pathogenic/likely pathogenic; VUS = variant of uncertain significance.

| **Rare Genetic Comparison** | **Composite** | **Hazard Ratio (95% CI)** | **Nominal p-value** |
| --- | --- | --- | --- |
| Thick vs. Thin | Primary | 1.92 (0.46-8.03) | 0.37 |
| MYBPC3 vs. MYH7 | Primary | 0.66 (0.33-1.33) | 0.24 |
| MYBPC3_pLOF vs. MYBPC3_nonLOF | Primary | 2.03 (0.74-5.53) | 0.45 |
| Sarcomere-P/LP vs. Sarcomere-VUS | Primary | 3.39 (0.81-14.2) | 0.10 |
| Thick vs. Thin | Secondary | 0.78 (0.48-1.27) | 0.32 |
| MYBPC3 vs. MYH7 | Secondary | 0.54 (0.38-0.78) | **0.000998** |
| MYBPC3_pLOF vs. MYBPC3_nonLOF | Secondary | 1.08 (0.64-1.82) | 0.77 |
| Sarcomere-P/LP vs. Sarcomere-VUS | Secondary | 0.99 (0.64-1.53) | 0.97 |

P-values derived via Wald test of Cox regression coefficient inequality from 0 and **bold** if nominally significant.

**Table 4.**
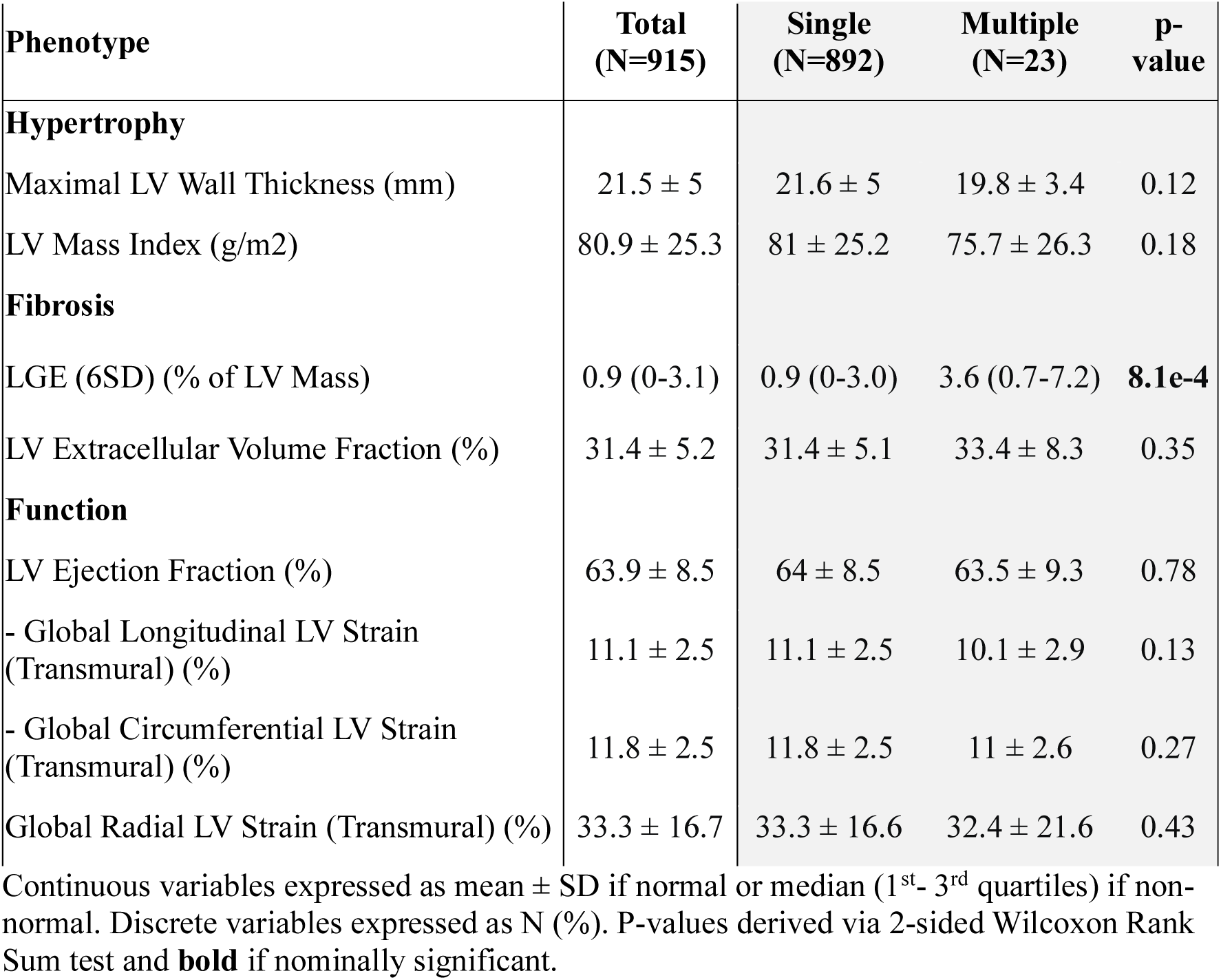
^4^Cardiac magnetic resonance imaging phenotypes of single and multiple-variant variant carriers in Hypertrophic Cardiomyopathy Registry. LGE = late gadolinium enhancement; LV = left ventricle

## Discussion

Previous studies have identified rare variant genotype-phenotype correlations in HCM^12,13,17,20^. Taken together however, they have presented mixed findings^16–20^, potentially reflecting limited sample sizes^12,13,16,19^, non-uniform phenotyping and variable recruitment biases. To overcome these limitations and definitively resolve discrepancies, we evaluated genotype-phenotype correlations in 915 sarcomere-positive patients of the NHLBI HCM Registry (Central Illustration). HCMR provides the largest registry of HCM patients with standardized phenotyping by CMR imaging – the gold-standard modality for phenotyping of cardiac structure and function^22^. Comparative studies have shown less inter-reader variability in measuring maxLVWT for CMR relative to echocardiography^37^ and an increased ability to detect mild hypertrophy^38^. This greater resolution may translate to increased detection of mild phenotypic differences between genotypic subgroups. CMR also measures replacement and interstitial fibrosis - notable prognostic phenotypes reflecting HCM severity^39,40^. As such, we leveraged HCMR’s standardized phenotyping of cardiac hypertrophy, fibrosis, and function, with clinical follow-up data, to identify robust associations between phenotypes and rare variant genotypes in HCM.

Specifically, we subgrouped patients using *a priori* approaches including gene-based, pathogenicity-based and count-based subgrouping and compared phenotypes across subgroups. Because hypertrophy, fibrosis and systolic function are routinely used for prognostication and management in HCM, differences across genotypic subgroups have direct clinical relevance. Unlike previous studies^17–21^, imaging-derived phenotype comparisons were adjusted for multiple testing and potential confounders including age and sex. For validation, we systematically reviewed prior studies and meta-analyzed findings. These showed that patients with disease-linked variants in thin-filament-encoding genes had less hypertrophy than those in thick-filament-encoding genes. This aligns with family studies of *TNNT2* and *TPM1* variants^12,13^, and cohort studies^17^. Thin-filament variants have also been associated with heart failure progression via LV systolic dysfunction^17,41^ but this was not observed in HCMR. LV hypertrophy is an established risk marker in HCM^42^ used in models such as HCM Risk-SCD^43^ and here, HCMR and prior analyses support reproducible and clinically interpretable differences in hypertrophy between these two genotypic subgroups.

Variants in the thick-filament-encoding genes *MYBPC3* and *MYH7* account for the majority of pathogenic HCM variants^2^. While some studies have identified phenotypic differences between *MYBPC3*-positive patients and *MYH7*-positive ones^19,20^, others have not^16,18^. In HCMR, the former subgroup had greater hypertrophy and lower LVEF. After meta-analysis with both supportive and contradictory prior studies, this remained significant, resolving prior discrepancies. In HCMR, *MYBPC3*-positive status was also associated with less incidence of the secondary composite, in particular atrial fibrillation, as previously observed in the Sarcomeric Human Cardiomyopathy Registry (SHaRe)^10^. SHaRe data also showed significantly greater risk of an HF composite and related events as transplantation/LVAD placement^10^. Together, these findings support clinically relevant phenotypic differences between *MYBPC3* and *MYH7* variant carriers for both prognostic markers as hypertrophy, and adverse clinical outcomes.

P/LP-positive and VUS-positive patients were also found to be phenotypically similar except for greater LGE in the former. This may in part be due to HCMR’s variant curation which subclassified VUSs into VUS_FP (favoured pathogenic), VUS, and VUS_FB (favoured benign). The VUSs described in this study exclude VUS_FBs and as such, may be biased towards pathogenicity. Nevertheless, previous cohort studies without such VUS subclassification also showed phenotypic similarities between the two subgroups, albeit with smaller sample sizes^21,33^. SHaRe data also indicated that hazard ratios for clinical outcomes were not significantly different (like in HCMR), with the exception of greater risk of severe systolic dysfunction (LVEF<35%) in P/LP-positive patients^10^. This study and previous ones (Supplemental Figure 6c) showed phenotypic similarities between the subgroups, suggesting that on average, such variants of ‘uncertain significance’ have some pathogenic effect and warrant attentive interpretation in clinic.

In HCMR, patients with multiple disease-linked variants were strikingly rare (∼1% of all patients) and did not differ significantly in phenotype from single-variant patients beyond elevated LGE. Previous work had suggested that 8% of gene-positive patients carried multiple P/LP variants but re-analysis of such studies highlighted a lack of rigorous pathogenicity classification and after reclassifying, indicated that the prevalence was instead 0.4%^36^. This aligns with our findings in HCMR (3/915 ≈ 0.3%) and suggests that predominately due to its rarity, carriage of multiple variants does not contribute much to variation in phenotypic severity in the general HCM population.

Overall, we leveraged the standardized CMR phenotyping with clinical follow-up, superior sample size, and genotypic diversity of HCMR to identify robust and significant genotype-phenotype correlations across genotype subgroups in sarcomere-positive HCM. Such associations are potentially clinically significant in that they map to routinely used markers of prognosis and management (hypertrophy, fibrosis, systolic function) and, for some, can be shown to influence outcome. This supports further evaluation of the role of rare variation as a factor that can refine prognosis and surveillance in the existing phenotype-centric framework.

### Study Limitations

A limitation of this work included the aggregation of numerous unique variants into per-gene groups. Given the substantial heterogeneity among different variants within the same gene^32,44,45^ (allelic heterogeneity), the gene-based subgrouping approach applied here averaged their varied effects. This greater within-group variation may have attenuated phenotypic differences between gene-based subgroups. Likewise, the modifying effects of additional factors such as common genetic variants^6^ and hypertension^7^ may have also affected between-group differences. Statistical limitations include the assumptions of multivariable linear and Cox regressions, and the limited number of clinical events in follow-up potentially leading to a lack of significant associations with adverse outcomes. This may reflect the low-risk nature of the HCMR cohort^11^. To increase statistical power, events were aggregated into composite outcomes. This came at the cost of understanding how variants relate to specific pathways of disease progression such as SCD or progressive HF. The rare nature of multiple-variant patients also limited statistical power to detect significant differences from single-variant patients. All sarcomere-positive patients were included in analyses regardless of kinship to maximise power. This may have introduced confounding effects, although the large majority of patients (∼92%^7^) were unrelated (<3 degrees of relatedness). The systematic review and meta-analyses were limited by 1) an inability to adjust for potential confounders because previous studies did not do so; 2) potential methodological CMR differences across studies such as the use of 6SD compared to visual cutoff for LGE quantification; 3) approximation of some prior studies’ medians and quartiles to means and standard deviations; 4) restriction to English-written literature.

## Conclusions

Previous studies of genotype-phenotype correlations have presented inconsistent findings. By leveraging the largest patient registry with rare-variant genotyping, standardized CMR phenotyping, and clinical follow-up, we identified and quantified such correlations, including greater hypertrophy in thick-filament variant-positive patients relative to thin-filament-positive ones. Of the former, patients with disease-linked variants in *MYBPC3* had greater hypertrophy and lower systolic function relative to those with *MYH7* variants, but a lower incidence of adverse clinical outcomes. Through systematic review and meta-analysis, we validated genotypic correlations with these prognostic markers and resolved prior discrepancies. We also showed that carriage of multiple disease-linked variants is rare in the unselected HCM population and consequently does not contribute much to variation in phenotypic severity. Future work may focus on disease driven by rare variants in genes beyond the core 8 sarcomeric genes such as *ALPK3*, thereby further resolving the genotype-phenotype correlation landscape in HCM.

## Data Availability

All data is in the manuscript and supplements.

## Acknowledgements

The authors acknowledge the technical guidance of Reem Malouf, MD, in meta-analysis, and the computational resources provided by the Biomedical Research Computing Facility at the University of Oxford. SN and HW acknowledge support from the Oxford NIHR Biomedical Research Centre.

## Abbreviations

CMR: cardiac magnetic resonance
ECVF: extracellular volume fraction
HCMR: Hypertrophic Cardiomyopathy Registry
LGE: late gadolinium enhancement
LVMi: indexed left ventricular mass
LVEF: left ventricular ejection fraction
maxLVWT: maximal left ventricular wall thickness
LoF: loss-of-function
P/LP: pathogenic/likely pathogenic
VUS: variant of uncertain significance

---

**Central Illustration.**
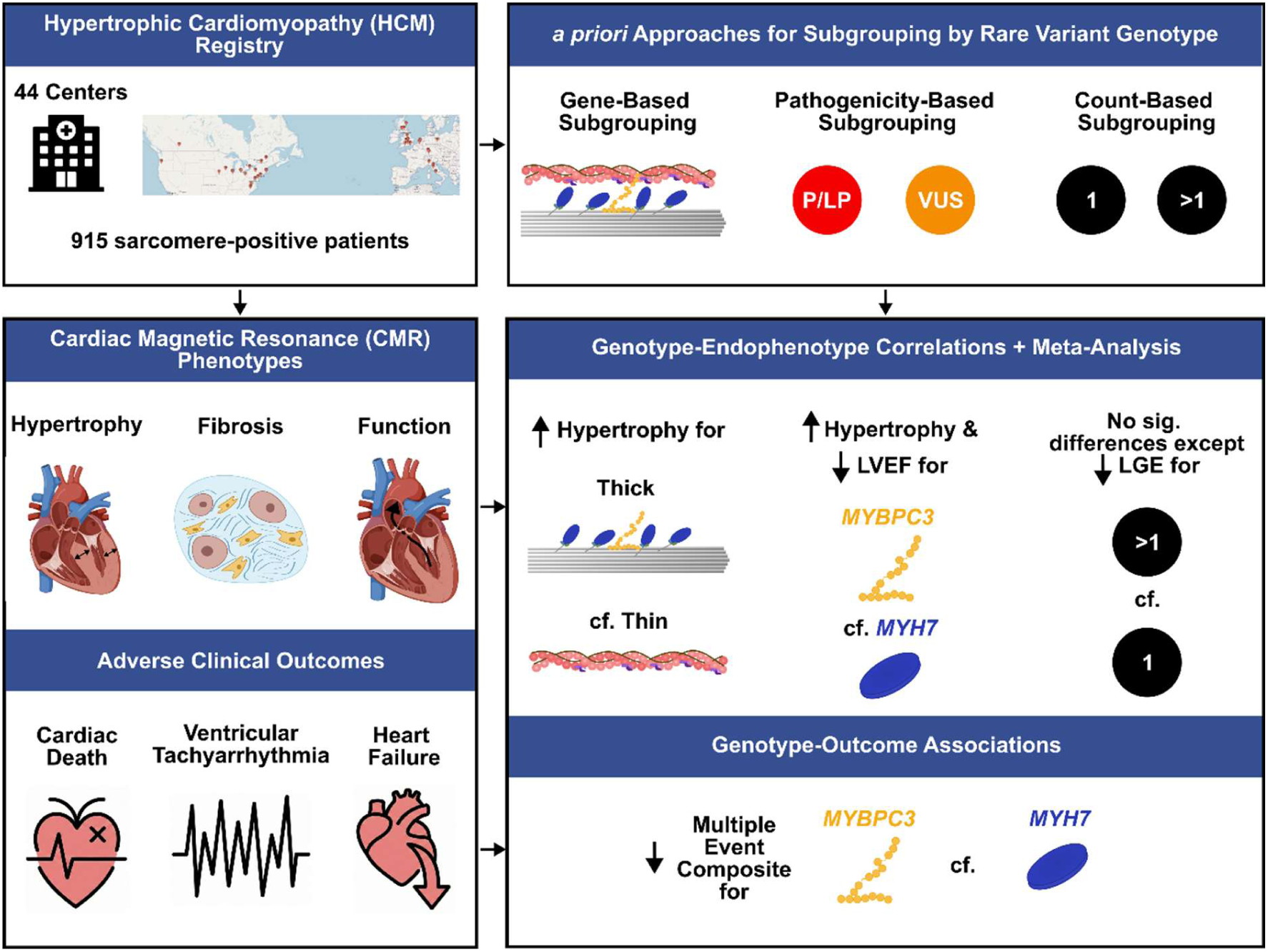
Genotype-phenotype correlations in sarcomeric variant-positive HCM. 915 sarcomeric variant-positive patients from 44 centres across North America and Europe in the National Heart, Lung, and Blood Institute Hypertrophic Cardiomyopathy Registry (HCMR) were subgrouped by genotype via three *a priori* approaches. Cardiac magnetic resonance imaging was used to measure left ventricular hypertrophy, fibrosis, and function. These were systematically compared across subgroups to identify genotype-phenotype correlations in the largest such study to date. Meta-analysis of primary analyses with prior studies identified by systematic review resolved past discrepancies and supported overall findings. Subgroups were also compared for association with adverse clinical outcomes to identify genotype-outcome associations. LGE = late gadolinium enhancement; LVEF = left ventricular ejection fraction; P/LP = Pathogenic/Likely pathogenic; VUS = variant of uncertain significance.

